# Multimodal Large Language Model Passes Specialty Board Examination and Surpasses Human Test-Taker Scores: A Comparative Analysis Examining the Stepwise Impact of Model Prompting Strategies on Performance

**DOI:** 10.1101/2024.07.27.24310809

**Authors:** Jamil S. Samaan, Samuel Margolis, Nitin Srinivasan, Apoorva Srinivasan, Yee Hui Yeo, Rajsavi Anand, Fadi S. Samaan, James Mirocha, Seyed Amir Ahmad Safavi-Naini, Bara El Kurdi, Ali Soroush, Rabindra Watson, Srinivas Gaddam, Joann G. Elmore, Brennan M.R. Spiegel, Nicholas P. Tatonetti

## Abstract

**Background:** Large language models (LLMs) have shown promise in answering medical licensing examination-style questions. However, there is limited research on the performance of multimodal LLMs on subspecialty medical examinations. Our study benchmarks the performance of multimodal LLM’s enhanced by model prompting strategies on gastroenterology subspeciality examination-style questions and examines how these prompting strategies incrementally improve overall performance.

**Methods:** We used the 2022 American College of Gastroenterology (ACG) self-assessment examination (N=300). This test is typically completed by gastroenterology fellows and established gastroenterologists preparing for the gastroenterology subspeciality board examination. We employed a sequential implementation of model prompting strategies: prompt engineering, retrieval augmented generation (RAG), five-shot learning, and an LLM-powered answer validation revision model (AVRM). GPT-4 and Gemini Pro were tested.

**Results:** Implementing all prompting strategies improved the overall score of GPT-4 from 60.3% to 80.7% and Gemini Pro’s from 48.0% to 54.3%. GPT-4’s score surpassed the 70% passing threshold and 75% average human test-taker scores unlike Gemini Pro. Stratification of questions by difficulty showed the accuracy of both LLMs mirrored that of human examinees, demonstrating higher accuracy as human test-taker accuracy increased. The addition of the AVRM to prompt, RAG and 5-shot increased GPT-4’s accuracy by 4.4%. The incremental addition of model prompting strategies improved accuracy for both non-image (57.2% to 80.4%) and image-based (63.0% to 80.9%) questions for GPT-4, but not Gemini Pro.

**Conclusions:** Our results underscore the value of model prompting strategies in improving LLM performance on subspecialty-level licensing exam questions. We also present a novel implementation of an LLM-powered reviewer model in the context of subspecialty medicine which further improved model performance when combined with other prompting strategies. Our findings highlight the potential future role of multimodal LLMs, particularly with the implementation of multiple model prompting strategies, as clinical decision support systems in subspecialty care for healthcare providers.

## INTRODUCTION

Large language models (LLMs) have shown potential to revolutionize medical information retrieval and clinical decision support.^1–4^ These advanced artificial intelligence (AI) systems leverage vast amounts of data and computational power to generate human-like responses.^5^ Previous investigations have demonstrated the accuracy and reliability of LLMs in answering clinical questions across various medical fields with promising results.^6–13^ As a more objective and generalizable benchmark for performance, studies have also explored LLMs’ impressive performance on standardized clinical examinations such as the United States Medical Licensing Examination (USMLE) and specialty examinations.^14–21^

Advancements in the capabilities of LLMs, such as image recognition, have opened new avenues for innovation and research into their potential applications in clinical care. Furthermore, model prompting strategies, such as prompt engineering, few-shot learning, and retrieval augmented generation (RAG) have provided promise in enhancing the performance of generalist foundation models on science and general medical knowledge benchmarks.^22–24^ Moreover, LLM self-reflection or verification has been shown to improve model accuracy, opening new avenues for the design of validation pipelines for model performance enhancement.^25–32^ There are currently no studies exploring the simultaneous implementation of these methods to enhance multimodal LLM performance in subspecialty medical examinations. Given the complex clinical reasoning required in specialty care, the role of model prompting strategies in LLM performance warrants further investigation.

The fields of gastroenterology and hepatology are ideal for examining multimodal LLM prompting strategies. These fields encompass a wide range of major topics and regularly utilize multiple diagnostic modalities, including cross-sectional imaging, endoscopy, ultrasound and manometry. A study in March 2023 evaluated the abilities of GPT-3.5 and GPT-4 to respond to simple prompts containing text-based questions from the 2021 and 2022 American College of Gastroenterology (ACG) Self-Assessment examinations.^33^ Neither model met the 70% passing threshold for these examinations. Although informative, the study had several limitations: it did not employ prompting strategies, predated the release of GPT-4 vision-LLMs, and relied on earlier iterations of both models. Another study investigating LLM performance on the ACG self-assessment examinations found significant enhancements using prompt engineering alone, although it was limited to only text-based LLMs and lacked incorporation of other model prompting strategies.^34^

Thus, we aimed to expand on these previous studies by examining the impact of model tuning on multimodal LLM performance on the 2022 ACG Self-Assessment examination to benchmark the performance of LLMs in gastroenterology and hepatology. To evaluate differences in LLM performance, we compared the performance of two widely used models, GPT-4 and Gemini Pro. To enhance model performance, we used the following model prompting strategies: prompt engineering, RAG, few-shot learning, and an LLM-powered peer reviewer termed Answer Validation Revision Model (AVRM). To better understand the impact of model prompting strategies, we analyzed the impact of stepwise implementation on overall performance.

## METHODS

### ACG Self-Assessment Examination

The ACG committee developed the ACG Self-Assessment to aid new and recertifying gastroenterologists for the American Board of Internal Medicine gastroenterology board examination.^35^ A new version is released annually, with the latest released in 2023. The exam consists of 300 multiple-choice questions covering core clinical knowledge topics in gastroenterology and hepatology, including medical images. Each question includes detailed explanations and scientific references. We used the 2023 version for parameter and prompt optimization and the 2022 version for testing.

### LLM Setup and Environment

We implemented model prompting strategies stepwise, starting with a zero-shot approach and sequentially adding strategies for a final combined approach (**Supplementary Figure 1)**. Given Gemini Pro’s poor performance, AVRM was not performed. Data were collected in April 2024, except for the AVRM model, designed and collected in May 2024. Refer to **Supplementary Methods** for further details

### Prompt Development

We iteratively developed and optimized a prompt using the 2023 ACG Self-Assessment questions. The prompt was refined based on a review of generated responses and overall performance. No gastroenterology or hepatology information was included; instead, we instructed the model on examination approaches, reasoning techniques, and comprehensive information evaluation.

### Retrieval Augmented Generation (RAG)

The RAG dataset included online publicly available clinical practice guidelines and practice updates from the ACG, American Gastroenterological Association (AGA), American Society for Gastrointestinal Endoscopy (ASGE), and American Association for the Study of Liver Diseases (AASLD). This approach resembles providing students with relevant reference materials as well as an effective and efficient method of identifying the most relevant sections for each question during an examination (**Figure 1**). Refer to **Supplementary Methods** for more details.

**Figure 1:**
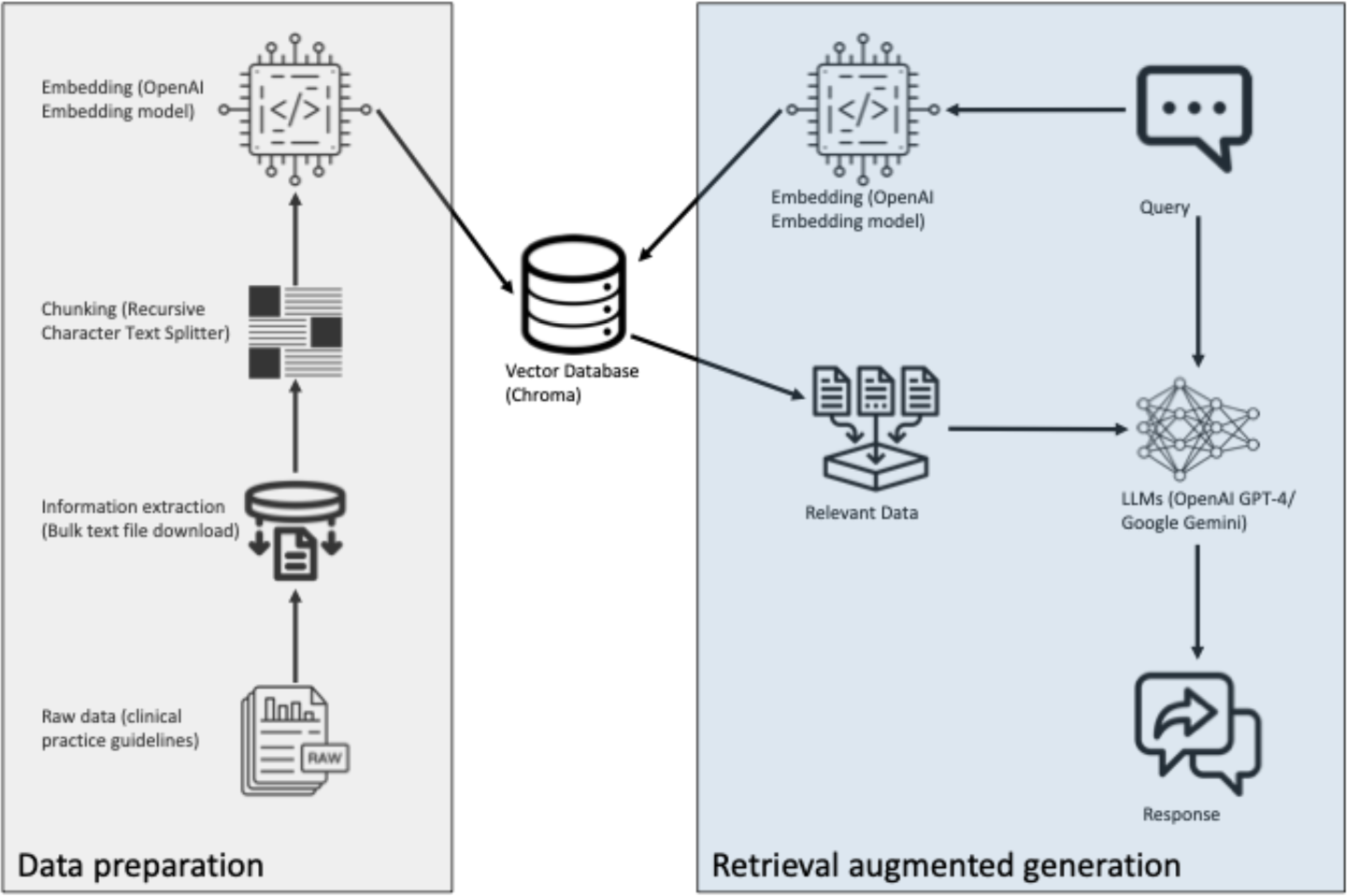
Diagram of Retrieval Augmented Generation Model Architecture for Question-Answering Using Attention-Based Deep Learning.

### *5-shot* Learning

We used a 5-shot learning strategy to enhance model performance. This is akin to students practicing for exams by completing practice questions. Each example included the question, answer choices, any associated images, and the explanation for the correct answer with annotations as provided by the ACG. Example selection was systematic and mirrored student preparation. During testing, we critically appraised the model’s performance on the ACG 2023 examination, prioritizing topics with more questions and lower scores to maximize improvement. We included straightforward guideline-based, and complex questions to ensure diverse examples. Lastly, we provided examples with and without images.

### Answer Validation Revision Model (AVRM)

The AVRM was implemented as the last step and tasked with first reviewing the inputs and outputs of the previous model, which utilized prompt, RAG and 5-shot, and then using the AVRM prompt to decide on the final answer to the question. An illustration of all inputs and outputs for the AVRM model are shown in **Supplementary Figure 2**. The AVRM prompt was developed by closely examining model reasoning and output on the 2023 ACG examination. AVRM prompt engineering included instructions to appraise the previous model’s output systematically. The AVRM ensured the previous model’s information summary was correct and logical. After examination, the AVRM either agreed with the previous model’s answer or disagreed and provided a new answer. We considered the answer ultimately chosen by the AVRM as the final answer for analysis. An example of model input and output is shown in **Supplementary Table 1**.

### Statistical Analysis

Two outcomes were considered: LLM accuracy and proportion of questions where LLMs chose the most popular answer chosen by human test-takers, irrespective of the correct answer. Multiple sub-analyses were conducted to better characterize LLM performance. Question difficulty, defined based on percentage of human test-takers choosing the correct answer, was categorized into 4 groups similar to prior study: least difficult (≥90%), mildly difficult (<90% and ≥75%), moderately difficult (≤75%), and very difficult (≤50%).^33^ Further sub-analyses included comparison of performance on image and non-image-based questions as well as difference in accuracy stratified by question topic as provided by the ACG. Further statistical analysis are described in **Supplemental Methods**.

To assess the impact of sequentially implementing model prompting strategies on LLM performance, we employed linear regression analyses. The explanatory variable was scaled 0-4 for GPT-4 and 0-3 for Gemini Pro as an ordinal variable with each increment representing an additional model prompting strategy. For the linear regression analysis, the data point zero-shot with prompt and RAG was excluded to ensure consistency in the progression of prompting interventions. The dependent variable was percentage of correct answers chosen or percent of most popular answers chosen by each LLM. An interaction term was used to examine the difference in slopes between Gemini Pro and GPT-4 as well as image and non-image-based questions. IBM SPSS Statistics version 29.0.2.0 was used for all analysis. The 0.05 significance level was used throughout.

## RESULTS

Our analysis included all 300 questions from the 2022 ACG Self-Assessment examination, with 138/300 (46%) containing images. At the final step of implementation, GPT-4’s performance improved from 60.3% to 80.7%, exceeding the 70% passing threshold and 75% average score of human test-takers (**Figure 2A**). Adding AVRM to prompt, RAG, and 5-shot increased GPT-4’s accuracy by 4.4%. At the final step of implementation, Gemini Pro’s performance improved from 48.0% to 54.3%, falling short of the minimum passing score (**Figure 2B**). GPT-4 (b=5.18, p<0.001) and Gemini Pro (b=1.99, p=0.024) both showed improvement with incremental prompting strategy implementation. GPT-4 improved at a 2.6-fold higher rate compared to Gemini Pro (p<0.001) (**Figure 2B**). Similar results were seen when examining trends in models choosing the most popular answer among human test-takers (**Supplementary Figure 3, Supplementary Figure 4**).

**Figure 2:**
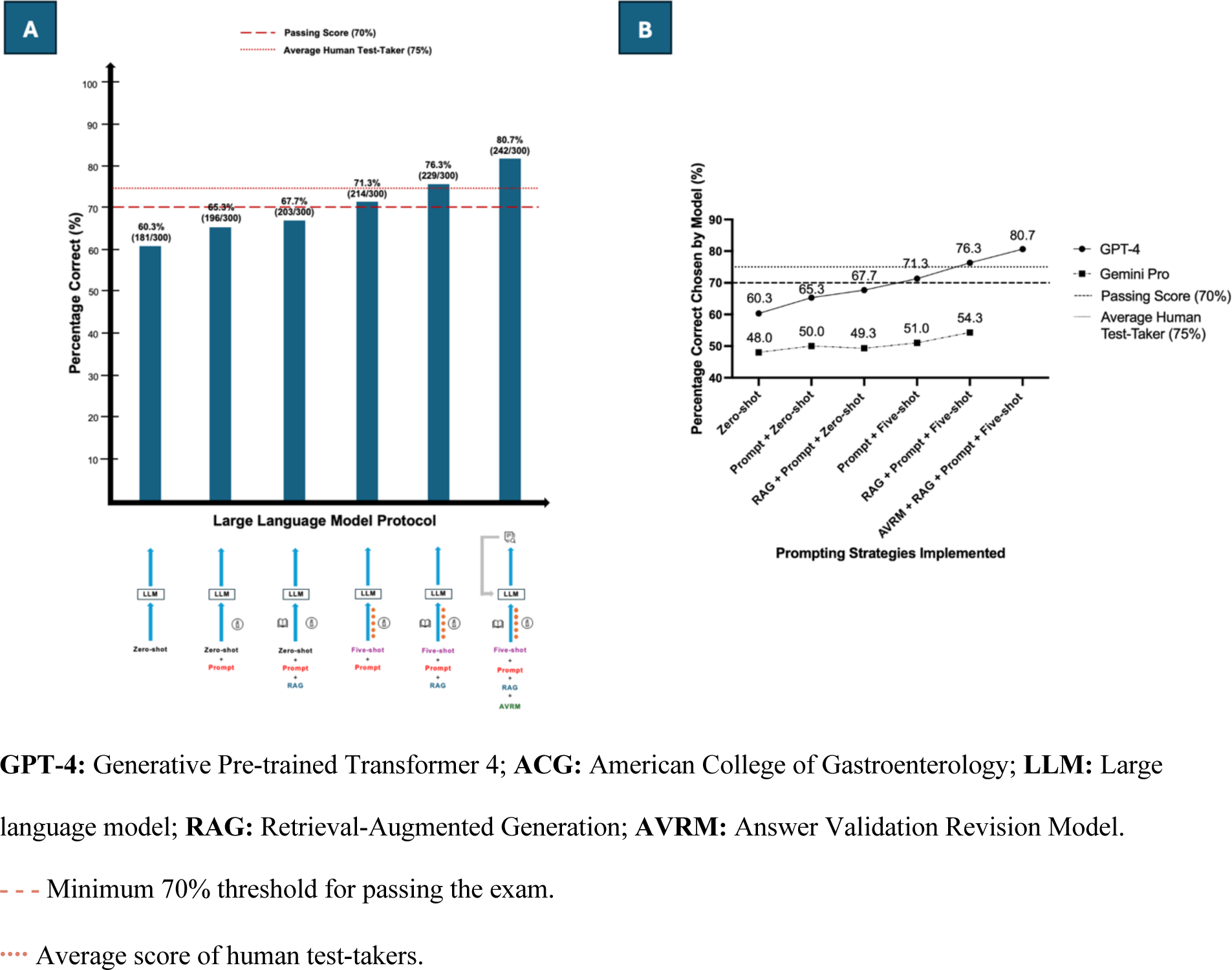
Stepwise Performance of GPT-4 (A) and performance comparison of GPT-4 and Gemini Pro (B) on the 2022 ACG Self-Assessment Examination with the Implementation of Each Model Prompting Strategy.

### LLM Performance Stratified by Image and Non-image Based Questions

GPT-4 showed worse zero-shot accuracy on image (57.3%) compared to non-image-based (63.0%) questions. Subsequently, GPT-4’s performance improved with the incremental addition of model prompting strategies on both non-image (b=4.45, p<0.001) and image (b=6.02, p<0.001) based questions. Performance for image-based questions improved at a greater rate compared to non-image-based questions (p=0.01), ultimately leading to a narrowing of the performance gap at the final step of implementation (image-based 80.4% vs non-image-based 80.9%) (**Figure 3A).** Gemini Pro did not demonstrate improvement in performance with the incremental addition of model prompting strategies on both non-image (b=3.03, p=0.10), and image (b=1.58, p=0.26) based questions without a difference in trends noted (p=0.176) (**Figure 3B**).

**Figure 3:**
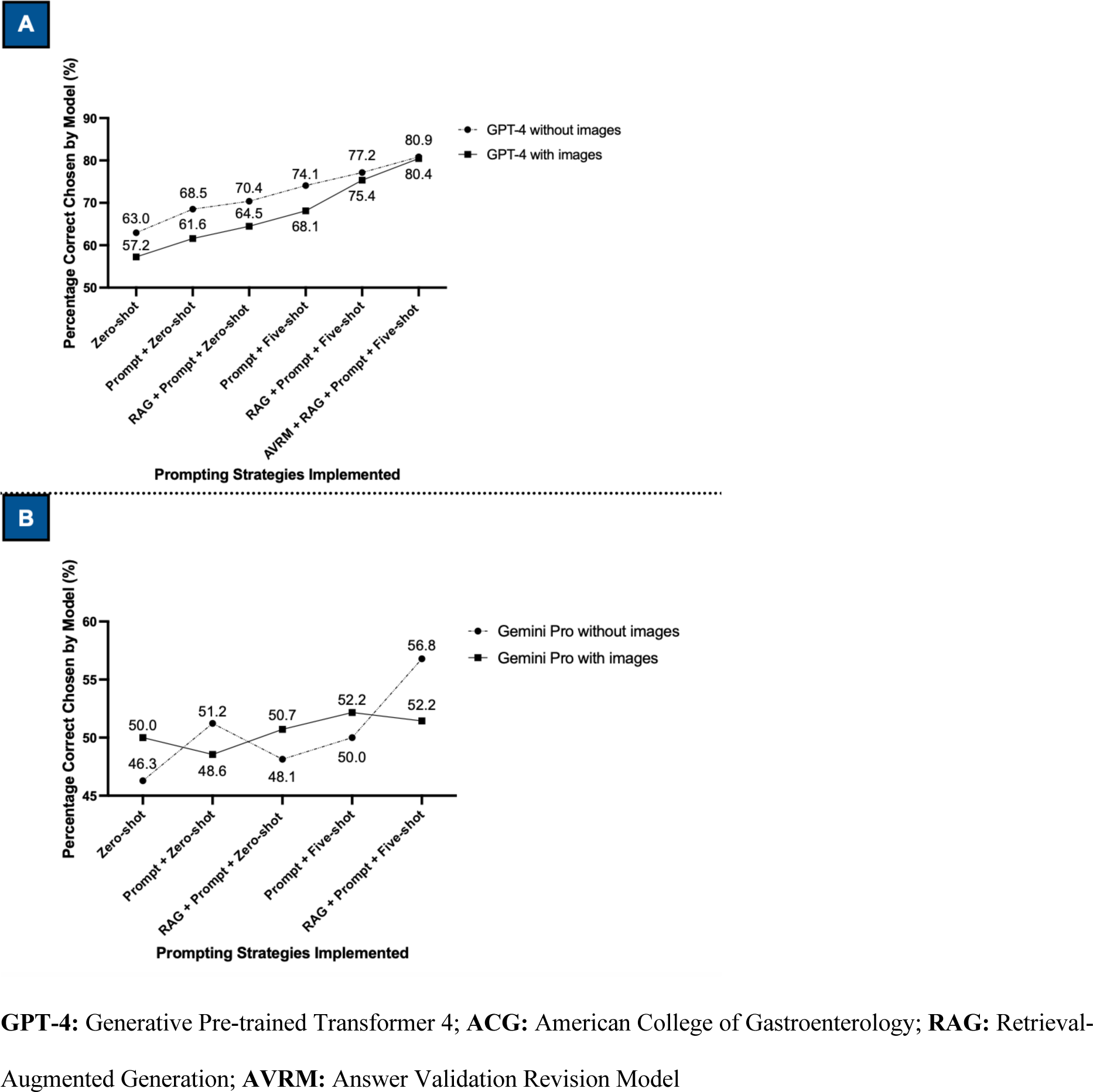
Stepwise Performance of GPT-4 (A) and Gemini Pro (B) on Image and Non-image containing 2022 ACG Self-Assessment Examination Questions with the Implementation of Each Model Prompting Strategy

### LLM Performance Stratified by Question Difficulty for Humans

A total of 75 questions were categorized as least difficult, 104 as mildly difficult, 119 as moderately difficult and 36 as very difficult. For both GPT-4 and Gemini Pro, accuracy increased with increasing human performance at all steps of implementation(**Figure 5, Supplemental Figure 5**). GPT-4’s performance improved with the incremental addition of model prompting strategies among all question difficulties: least difficult (b=4.4, p=0.039), mildly difficult (b=3.95, p<0.001), moderately difficult (b=6.65, p=0.003), very difficult (b=2.78, p=0.046) (**Figure 4**). Gemini Pro demonstrated improvement in performance among only the hardest questions (**Supplemental Figure 5**).

**Figure 4:**
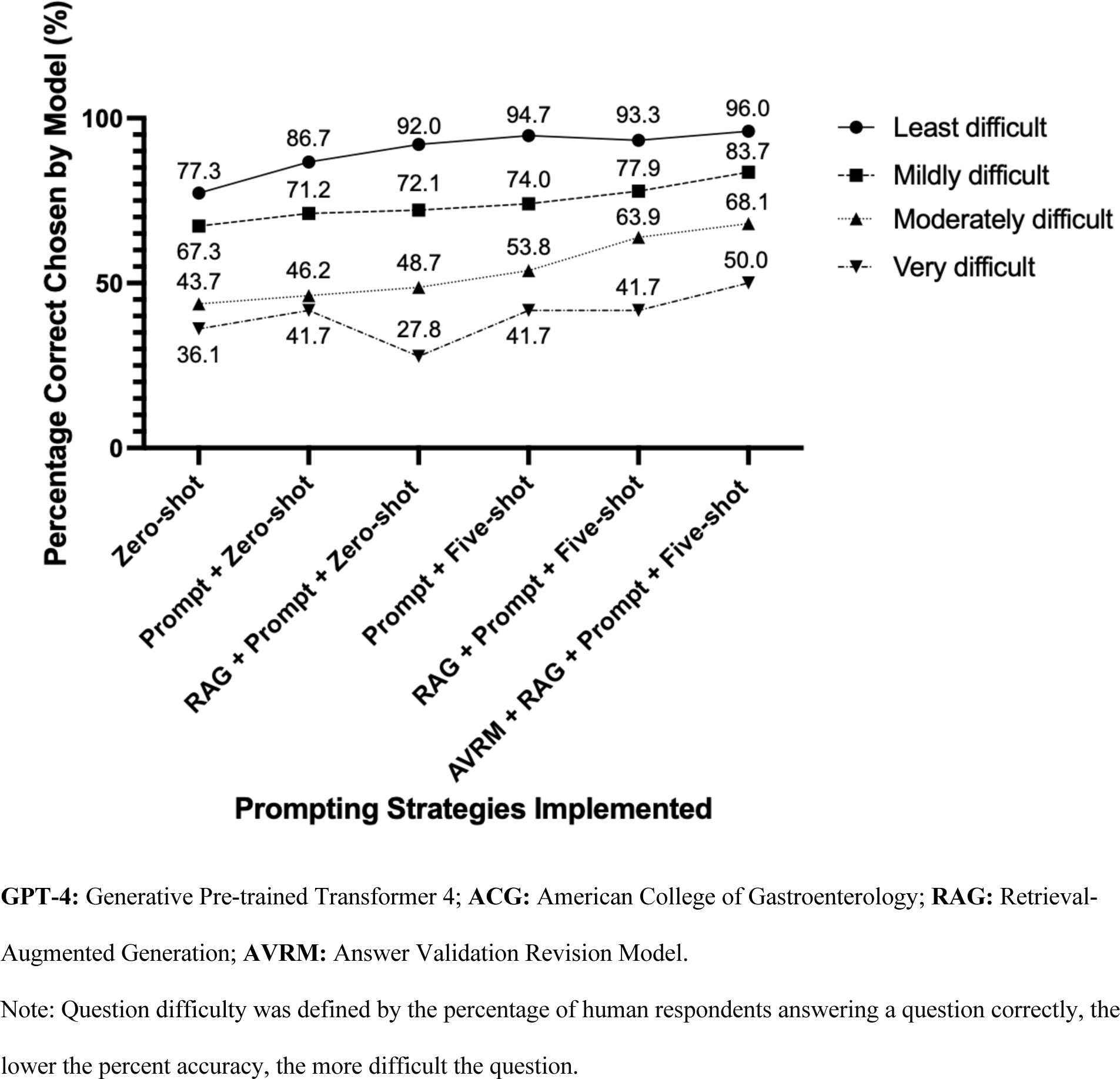
Stepwise Performance of GPT-4 on the 2022 ACG Self-Assessment Examination with the Implementation of Each Model Prompting Strategy Stratified by Question Difficulty.

**Figure 5:**
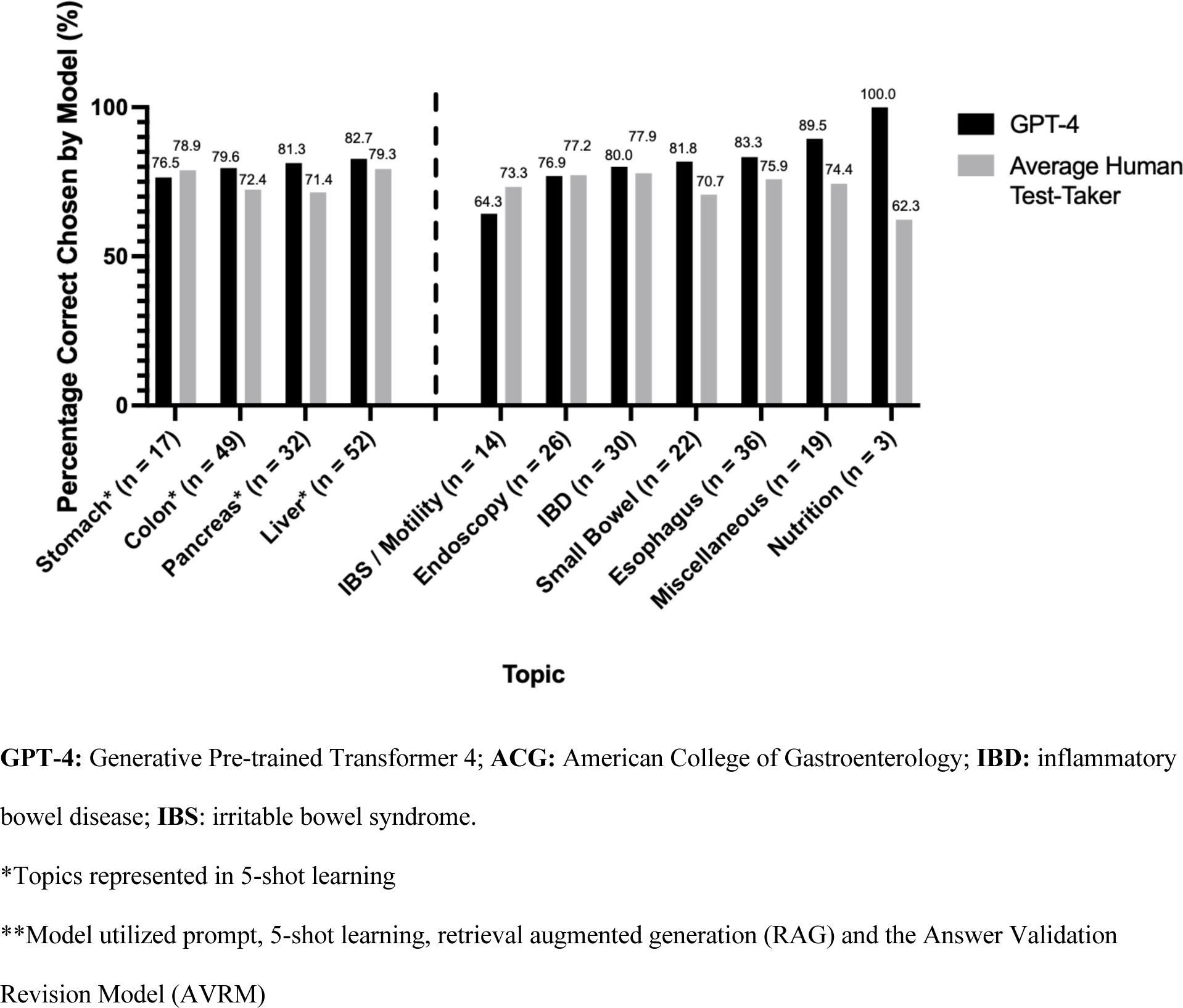
Stepwise Performance of GPT-4 at the Final Step of Model Prompting Strategy Implementation** on the 2022 ACG Self-Assessment Examination Questions with the Implementation of Each Model Prompting Strategy Stratified by Topic.

### LLM Performance Stratified by Question Topic

GPT-4 performed best on the topics Miscellaneous (17/19, 89.5%) and performed worst on the topic irritable bowel syndrome (IBS)/motility (9/14, 64.3%) (**Figure 5**). Gemini Pro performed best on IBS/motility (11/14,78.6%) and worst on Colon (12/49, 24.5%) (**Supplemental Figure 6**). The most difficult topic was Nutrition (62.3%) while the easiest was Liver (79.3%). There was no significant difference in the change in accuracy when comparing prompt and zero-shot and prompt and 5-shot for topics included (median=-4.9, IQR=-15.2, 8.4) and not included (median=4.5, IQR=0, 13.9) in 5-shot learning for GPT-4 (p=0.11). Similar results were seen for Gemini Pro for topics included (median=-2.95, IQR=-6.1, 8.6) and not included (median=0, IQR=-30.8, 5.6) (p=0.78).

## DISCUSSION

We examined the stepwise impact of stepwise implementation of prompting strategies on multimodal LLM performance, demonstrating for the first time an LLM passing the ACG Self-Assessment examination. Both GPT-4 and Gemini Pro showed performance enhancements with successive implementation of prompting strategies. However, GPT-4 outperformed Gemini Pro on all metrics at all steps of implementation. At the final step of prompting strategy implementation GPT-4 scored 80.7%, surpassing the 70% passing threshold and the 75% average score of human test-takers. This analysis establishes a new benchmark for multimodal LLM performance in gastroenterology and hepatology, highlighting the benefits of simultaneous implementation of multiple prompting strategies.

GPT-4 consistently outperformed Gemini Pro across all levels of prompting strategy implementation. These results corroborate prior studies demonstrating GPT-4’s superior performance over Gemini Pro in domains such as ophthalmology surgical planning, hypertension education, and glaucoma management.^36–38^ Its also notable that GPT-4 showed higher rate of performance improvement with successive implementation of model prompting strategies compared to Gemini Pro. The discrepancy in performance is likely multifactorial, arising from variations in model architecture, training data, and post-training fine-tuning. This performance gap underscores the importance of model selection in developing high-performing clinical decision support systems.

GPT-4 and Gemini Pro accuracy mimicked that of human test takers, as the models were more likely to select a correct answer when humans were more likely to answer correctly. This finding was noted at every step of implementation for both models (**Figure 4, Supplemental Figure 5**). While not directly investigated by our analysis, there are multiple potential reasons for this trend that are worth exploring in future studies. Multiple factors can contribute to question difficulty such as requirement of higher order thinking, content familiarity, question format and question clarity. LLMs have been previously shown to mirror patterns of human reasoning, demonstrating many of the same qualitative human patterns of logical reasoning tasks.^39^ Importantly, these trends were seen across all LLMs tested and all training and tuning paradigms, suggesting a general phenomenon among LLMs.^39^ Its plausible that the trends observed in our analysis are in part due to the models aligning with human decision-making patterns. Perhaps LLMs utilize common reasoning pathways or heuristics that humans employ when faced with similar questions. Future investigations into LLM reasoning patterns compared to humans in the context of clinical reasoning are warranted.

Similar model performance was seen among topics included and not included in few-shot learning when comparing zero shot and prompt with 5-shot and prompt performance (**Figure 5, Supplemental Figure 6**). These results imply that topic selection may not be a limiting factor in future studies involving a few-shot learning design for certain LLMs. We recommend emphasis on other criteria utilized in our study, such as question complexity and training set performance as described in our methods.

GPT-4 and Gemini Pro performed better on some topics compared to others (**Figure 5**, **Supplemental Figure 6**). GPT-4 notably performed poorly on the IBS/motility questions, while Gemini Pro performed best on this topic. The exam questions for IBS/motility were of average difficulty when examining average human test-taker scores on all topics (**Figure 5**), making disproportionately more difficult questions for IBS/motility a less likely explanation. The reason for this discrepancy in performance may be due to difference in training data source and fine-tuning although this warrants further investigation in future studies.

Previous studies have examined the ability of LLMs to self-reflect and improve performance with promising results. The concept of self-reflection entails LLMs assessing their own output, or that of other LLMs, to improve reasoning and ultimately accuracy. Studies in the literature used a variety of designs utilizing different prompting strategies, incorporation of external information, few-shot learning, RAG and the utilization of multiple LLMs as self-reflection agents.^25–32^ The AVRM in our study is a hybrid of previously described methods with certain variations that are specific to our study design such as the simultaneous incorporation of 5-shot learning and RAG as AVRM input as well as including a gastroenterology and hepatology guideline library for the RAG. While our findings are promising there are multiple opportunities for optimizing the AVRM performance in the context of subspecialty care. One consideration is optimizing AVRM inputs to maximize performance. This includes optimizing the AVRM prompt, determining the ideal quantity and types of examples for few-shot learning, and fine-tuning the source content and parameters of the Retrieval-Augmented Generation (RAG) framework. Additionally, improvements in the design of the AVRM pipeline warrant consideration. This includes optimizing the sequence of input processing and evaluating the number and types of LLMs required to achieve optimal performance.

GPT-4’s disparity in performance between image and non-image-based questions noticeably narrowed after the implementation of prompting, RAG, and 5-shot and nullified after the further implementation of AVRM. The lack of impact of 5-shot learning and prompting alone on the performance gap was unexpected, given that some examples contained images. Furthermore, the addition of RAG to prompting and 5-shot learning, which did not include images, appears to have disproportionately improved the models’ performance on image-based questions. The improvement seen with the addition of AVRM was expected and potentially highlights the utility of an additional layer of decision-making that specifically evaluates and potentially corrects responses generated by the primary model. However, the reason for the disproportionate impact of this added benefit on image-based questions is unclear and warrants future investigation.

### Limitations

Our study has several limitations to consider. We ran one exam for each step of implementation making examining consistency in accuracy not possible. While board examinations serve as an objective benchmark for LLM performance, board questions do not represent the complexity of all real-world clinical scenarios, and future prospective studies are required to assess LLM performance in clinical decision-making more comprehensively. The source of training data for both LLMs has not been publicly disclosed, making critical appraisal of the sources used to train the models not possible. Our RAG was constructed in plain text format which fails to capture information in pictures and graphs. While language in charts were represented in the RAG, chart formation and spacing was not preserved making nuanced interpretation of the information difficult. Furthermore, our RAG library only contained society guidelines which are not comprehensive of all topics and knowledge in the fields of gastroenterology and hepatology. Future studies incorporating images in guidelines as well as including other relevant source material may be helpful in improving model performance. Our study utilized linear regression analysis to evaluate trends and differences in trends, with prompting strategy implementation (the explanatory variable) treated as an ordinal variable, while standard linear regression analysis assumes that a numerical explanatory variable is measured on an equal interval level. The intent of our analysis was exploratory, and the results should be interpreted as suggestive only. Future studies are needed to evaluate the statistical significance of these trends and their differences more conclusively. Lastly, cost effectiveness and token usage were not explored in our study and warrants future examination to better understand the economic impact and practicality of LLMs as decision support tools.

## CONCLUSION

GPT-4, with the simultaneous utilization of model prompting strategies, surpassed both the passing threshold and the average human test-taker’s scores on the ACG Self-Assessment examination. GPT-4 outperformed Gemini Pro at all levels of implementation including a greater rate of improvement in performance with sequential implementation of strategies. We also show improvement in overall performance by implementing an LLM-powered reviewer model. Our analysis underscores the significant improvement in LLM standardized subspecialty examination performance. Moreover, the performance of GPT-4 enhanced by model prompting strategies underscores the promising future role of LLMs as adjunctive tools in clinical decision support, augmenting the care delivered by healthcare providers. In view of the complex and nuanced nature of clinical medicine, we support the future use of LLMs to supplement, rather than supplant, the care provided by licensed healthcare professionals. Prospective studies are needed to better understand the performance of LLMs using model prompting strategies in real-world complex medical decision-making settings.

## Data Availability

2022 American College of Gastroenterology (ACG) self-assessment examination. Available at https://education.gi.org/satest/satest_18

## Declaration of AI and AI-assisted technologies in the writing process

During the preparation of this work, the authors used GPT-4 to improve readability and language. After using this tool/service, the authors reviewed and edited the content as needed and take full responsibility for the content of the publication.

## Data and code availability

American College of Gastroenterology examination questions are available at https://members.gi.org.

## Author Contributions

J.S.S.: Conceptualization, co-designed the analyses, co-designed models tested, performed data collection, performed statistical analysis, co-wrote the manuscript, and approved the final draft. S.M.: Co-designed the analyses, co-designed models tested, built the models tested, performed data collection, co-wrote the manuscript, and approved the final draft.

N.S.: Performed data collection, edited the manuscript for important intellectual content, and approved the final draft.

A.S.: Performed data collection, edited the manuscript for important intellectual content, and approved the final draft.

Y.Y: Performed statistical analysis, edited the manuscript for important intellectual content and approved the final draft.

R.A.: Performed data collection, edited the manuscript for important intellectual content, and approved the final draft.

F.S.S.: Performed data collection, edited the manuscript for important intellectual content and approved the final draft.

J.M.: Performed statistical analysis, edited the manuscript for important intellectual content and approved the final draft.

S.A.A.S.N.: Edited the manuscript for important intellectual content and approved the final draft. B.E.K.: Edited the manuscript for important intellectual content and approved the final draft.

A.S.: Edited the manuscript for important intellectual content and approved the final draft.

R.W.: Edited the manuscript for important intellectual content and approved the final draft.

S.G.: Edited the manuscript for important intellectual content and approved the final draft.

J.G.E.: Edited the manuscript for important intellectual content and approved the final draft.

B.M.R.S.: Edited the manuscript for important intellectual content and approved the final draft.

N.P.T.: Supervision, edited the manuscript for important intellectual content, and approved the final draft.

## Abbreviations

ChatGPT: Chat Generative Pre-trained Transformer
LLM: Large language model
AI: Artificial Intelligence
USMLE: United States Medical Licensing Examination
RAG: Retrieval Augmented Generation
AGA: American Gastroenterological Association
ASGE: American Society for Gastrointestinal Endoscopy
AASLD: American Association for the Study of Liver Diseases
AVRM: Answer Validation Revision Model

## REFERENCES

1. Benary M, Wang XD, Schmidt M, et al. Leveraging Large Language Models for Decision Support in Personalized Oncology. JAMA Netw Open. 2023;6(11):e2343689. doi:10.1001/jamanetworkopen.2023.43689

2. Gilbert S, Kather JN, Hogan A. Augmented non-hallucinating large language models as medical information curators. npj Digit Med. 2024;7(1):100. doi:10.1038/s41746-024-01081-0

3. Perlis RH, Goldberg JF, Ostacher MJ, Schneck CD. Clinical decision support for bipolar depression using large language models. Neuropsychopharmacol. Published online March 13, 2024. doi:10.1038/s41386-024-01841-2

4. Shahab O, El Kurdi B, Shaukat A, Nadkarni G, Soroush A. Large language models: a primer and gastroenterology applications. Therap Adv Gastroenterol. 2024;17:17562848241227031. doi:10.1177/17562848241227031

5. OpenAI. GPT-4 Technical Report. Published online 2023. doi:10.48550/ARXIV.2303.08774

6. Samaan JS, Yeo YH, Rajeev N, et al. Assessing the Accuracy of Responses by the Language Model ChatGPT to Questions Regarding Bariatric Surgery. OBES SURG. Published online April 27, 2023. doi:10.1007/s11695-023-06603-5

7. Yeo YH, Samaan JS, Ng WH, et al. Assessing the performance of ChatGPT in answering questions regarding cirrhosis and hepatocellular carcinoma. Clin Mol Hepatol. Published online March 22, 2023. doi:10.3350/cmh.2023.0089

8. Ayers JW, Poliak A, Dredze M, et al. Comparing Physician and Artificial Intelligence Chatbot Responses to Patient Questions Posted to a Public Social Media Forum. JAMA Intern Med. 2023;183(6):589. doi:10.1001/jamainternmed.2023.1838

9. Sciberras M, Farrugia Y, Gordon H, et al. Accuracy of Information given by ChatGPT for Patients with Inflammatory Bowel Disease in Relation to ECCO Guidelines. Journal of Crohn’s and Colitis. Published online March 23, 2024:jjae040. doi:10.1093/ecco-jcc/jjae040

10. Klang E, Sourosh A, Nadkarni GN, Sharif K, Lahat A. Evaluating the role of ChatGPT in gastroenterology: a comprehensive systematic review of applications, benefits, and limitations. Therap Adv Gastroenterol. 2023;16:17562848231218618. doi:10.1177/17562848231218618

11. Han T, Adams LC, Bressem KK, Busch F, Nebelung S, Truhn D. Comparative Analysis of Multimodal Large Language Model Performance on Clinical Vignette Questions. JAMA. 2024;331(15):1320. doi:10.1001/jama.2023.27861

12. Zakka C, Shad R, Chaurasia A, et al. Almanac — Retrieval-Augmented Language Models for Clinical Medicine. NEJM AI. 2024;1(2). doi:10.1056/AIoa2300068

13. Saab K, Tu T, Weng WH, et al. Capabilities of Gemini Models in Medicine. Published online May 1, 2024. Accessed July 16, 2024. http://arxiv.org/abs/2404.18416

14. Gilson A, Safranek CW, Huang T, et al. How Does ChatGPT Perform on the United States Medical Licensing Examination? The Implications of Large Language Models for Medical Education and Knowledge Assessment. JMIR Med Educ. 2023;9:e45312. doi:10.2196/45312

15. Shieh A, Tran B, He G, Kumar M, Freed JA, Majety P. Assessing ChatGPT 4.0’s test performance and clinical diagnostic accuracy on USMLE STEP 2 CK and clinical case reports. Sci Rep. 2024;14(1):9330. doi:10.1038/s41598-024-58760-x

16. Sumbal A, Sumbal R, Amir A. Can ChatGPT-3.5 Pass a Medical Exam? A Systematic Review of ChatGPT’s Performance in Academic Testing. Journal of Medical Education and Curricular Development. 2024;11:23821205241238641. doi:10.1177/23821205241238641

17. Van Nuland M, Erdogan A, Aςar C, et al. Performance of ChatGPT on Factual Knowledge Questions Regarding Clinical Pharmacy. The Journal of Clinical Pharma. Published online April 16, 2024:jcph.2443. doi:10.1002/jcph.2443

18. Botross M, Mohammadi SO, Montgomery K, Crawford C. Performance of Google’s Artificial Intelligence Chatbot “Bard” (Now “Gemini”) on Ophthalmology Board Exam Practice Questions. Cureus. Published online March 31, 2024. doi:10.7759/cureus.57348

19. Alexandrou M, Mahtani AU, Rempakos A, et al. Performance of ChatGPT on ACC/SCAI Interventional Cardiology Certification Simulation Exam. JACC: Cardiovascular Interventions. Published online May 2024:S1936879824005740. doi:10.1016/j.jcin.2024.03.012

20. D’Anna G, Van Cauter S, Thurnher M, Van Goethem J, Haller S. Can large language models pass official high-grade exams of the European Society of Neuroradiology courses? A direct comparison between OpenAI chatGPT 3.5, OpenAI GPT4 and Google Bard. Neuroradiology. Published online May 6, 2024. doi:10.1007/s00234-024-03371-6

21. Wu S, Koo M, Blum L, et al. Benchmarking Open-Source Large Language Models, GPT-4 and Claude 2 on Multiple-Choice Questions in Nephrology. NEJM AI. 2024;1(2). doi:10.1056/AIdbp2300092

22. Nori H, Lee YT, Zhang S, et al. Can Generalist Foundation Models Outcompete Special-Purpose Tuning? Case Study in Medicine. Published online 2023. doi:10.48550/ARXIV.2311.16452

23. Liu F, Zhu T, Wu X, et al. A medical multimodal large language model for future pandemics. npj Digit Med. 2023;6(1):226. doi:10.1038/s41746-023-00952-2

24. Meskó B. The Impact of Multimodal Large Language Models on Health Care’s Future. J Med Internet Res. 2023;25:e52865. doi:10.2196/52865

25. Ling Z, Fang Y, Li X, et al. Deductive Verification of Chain-of-Thought Reasoning. Published online October 3, 2023. Accessed July 23, 2024. http://arxiv.org/abs/2306.03872

26. Peng B, Galley M, He P, et al. Check Your Facts and Try Again: Improving Large Language Models with External Knowledge and Automated Feedback. Published online March 8, 2023. Accessed July 23, 2024. http://arxiv.org/abs/2302.12813

27. Gero Z, Singh C, Cheng H, et al. Self-Verification Improves Few-Shot Clinical Information Extraction. Published online May 30, 2023. Accessed July 23, 2024. http://arxiv.org/abs/2306.00024

28. An S, Ma Z, Lin Z, Zheng N, Lou JG, Chen W. Learning From Mistakes Makes LLM Better Reasoner. Published online March 29, 2024. Accessed July 23, 2024. http://arxiv.org/abs/2310.20689

29. Madaan A, Tandon N, Gupta P, et al. Self-Refine: Iterative Refinement with Self-Feedback. Published online May 25, 2023. Accessed July 23, 2024. http://arxiv.org/abs/2303.17651

30. Pan L, Saxon M, Xu W, Nathani D, Wang X, Wang WY. Automatically Correcting Large Language Models: Surveying the landscape of diverse self-correction strategies. Published online August 29, 2023. Accessed July 23, 2024. http://arxiv.org/abs/2308.03188

31. Jeong M, Sohn J, Sung M, Kang J. Improving medical reasoning through retrieval and self-reflection with retrieval-augmented large language models. Bioinformatics. 2024;40(Supplement_1):i119-i129. doi:10.1093/bioinformatics/btae238

32. Renze M, Guven E. Self-Reflection in LLM Agents: Effects on Problem-Solving Performance. Published online May 5, 2024. Accessed July 18, 2024. http://arxiv.org/abs/2405.06682

33. Suchman K, Garg S, Trindade AJ. Chat Generative Pretrained Transformer Fails the Multiple-Choice American College of Gastroenterology Self-Assessment Test. Am J Gastroenterol. 2023;118(12):2280–2282. doi:10.14309/ajg.0000000000002320

34. Ali S, Shahab O, Al Shabeeb R, et al. General purpose large language models match human performance on gastroenterology board exam self-assessments. Published online September 25, 2023. doi:10.1101/2023.09.21.23295918

35. Self-Assessment Test. Accessed May 5, 2024. https://members.gi.org/store/listitems2.asp?type=1&prodcat=11&cn-reloaded=1

36. Stanaway JD, Afshin A, Gakidou E, et al. Global, regional, and national comparative risk assessment of 84 behavioural, environmental and occupational, and metabolic risks or clusters of risks for 195 countries and territories, 1990–2017: a systematic analysis for the Global Burden of Disease Study 2017. The Lancet. 2018;392(10159):1923-1994. doi:10.1016/S0140-6736(18)32225-6

37. Lee TJ, Campbell DJ, Patel S, et al. Unlocking Health Literacy: The Ultimate Guide to Hypertension Education From ChatGPT Versus Google Gemini. Cureus. Published online May 8, 2024. doi:10.7759/cureus.59898

38. Carlà MM, Gambini G, Baldascino A, et al. Large language models as assistance for glaucoma surgical cases: a ChatGPT vs. Google Gemini comparison. Graefes Arch Clin Exp Ophthalmol. Published online April 4, 2024. doi:10.1007/s00417-024-06470-5

39. Lampinen AK, Dasgupta I, Chan SCY, et al. Language models, like humans, show content effects on reasoning tasks. Abbott D, ed. PNAS Nexus. 2024;3(7):pgae233. doi:10.1093/pnasnexus/pgae233

