## Supplementary Figure 1-6; Supplementary Methods, Supplementary Table 1 for "Multimodal Large Language Model Passes Specialty Board Examination and Surpasses Human Test-Taker Scores: A Comparative Analysis Examining the Stepwise Impact of Model Prompting Strategies on Performance"

### Supplemental Materials

Table of Contents:


1. Supplemental Figure 1………………………………………………………………………………………. Page 2
2. Supplementary Methods………………………………………………………………………………Page 3-4
3. Supplemental Figure 2…………………………………………………………………………………………Page 5
4. Supplementary Table 1……………………………………………………………………………………Pages 6-7
5. Supplemental Figure 3………………………………………………………………………………………..Page 8-9
6. Supplemental Figure 4……………………………………………………………………………………Page 10-11
7. Supplemental Figure 5………………………………………………………………………………………Page 12
8. Supplemental Figure 6………………………………………………………………………………………Page 13
9. References…………………………………………………………………………………………………………Page 14

**Supplementary Figure 1**: Illustration of GPT-4 and Gemini Pro Step-Wise Response Generation to Questions from the 2022 American College of Gastroenterology (ACG) Self Assessment.

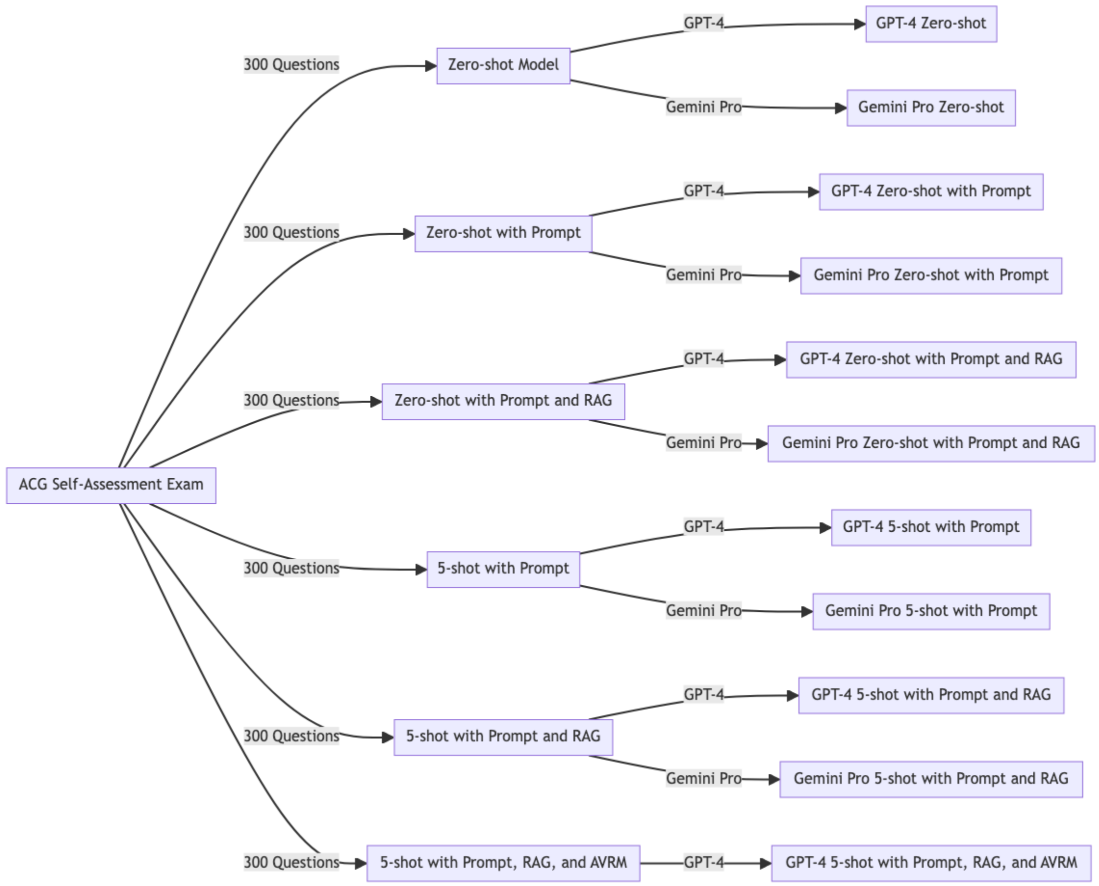


**GPT-4:** Generative Pre-trained Transformer 4; **RAG:** Retrieval-Augmented Generation; **AVRM:** Answer Validation Revision Model.

#### Supplementary Methods

##### *LLM Setup and Environment*

Each question was tested in a new session. GPT-4-turbo-0301 and gemini-1.0-pro-vision-001 were used. The LLMs were accessed through the application programming interface (API). The API call included the overall reviewer prompt, the user prompt, the question, GPT-4 Turbo, and a batch of images. The temperature was set to 1 and maximum token 4,096. Temperature is the default value and max tokens was capped to maintain costs and consistency.

##### *Retrieval Augmented Generation (RAG)*

Text within tabular and visual data were encoded as plain text format. We used a Recursive Character Text Splitter designed to segment text into chunks of 500 characters with an overlap of 50 characters between consecutive chunks.[1] This method ensures continuity and context retention across segments, enhancing the performance of downstream natural language processing tasks. To embed the data, we utilized the OpenAI embedding model text-embedding-ada-002 to transform text into high-dimensional vectors that capture semantic meaning, allowing for efficient comparison and retrieval of similar content.[2] By encoding textual data into these embeddings, we could more efficiently query the database to pull relevant text chunks for the model during response generation. For each examination question presented to the LLM, the RAG model selected the four most relevant text chunks as additional input.

##### *Statistical Analysis*

We present descriptive analyses as counts and percentages. Human performance was defined as the mean accuracy for all human test-takers as provided by the ACG. Colon, Liver, Stomach, and Pancreas were represented in the 5-shot learning examples. To ascertain any disproportionate impact of including 4 out of 11 topics in the 5-shot examples on overall performance, we evaluated changes in LLM accuracy between prompt and zero-shot compared to prompt and 5-shot for each topic. Descriptive statistics for this analysis are presented as median and interquartile ranges. Difference in change in performance was evaluated using the Mann-Whitney U Test.

**Supplementary Figure 2**: Illustration of Inputs and Outputs for the Answer Validation Revision Model (AVRM).

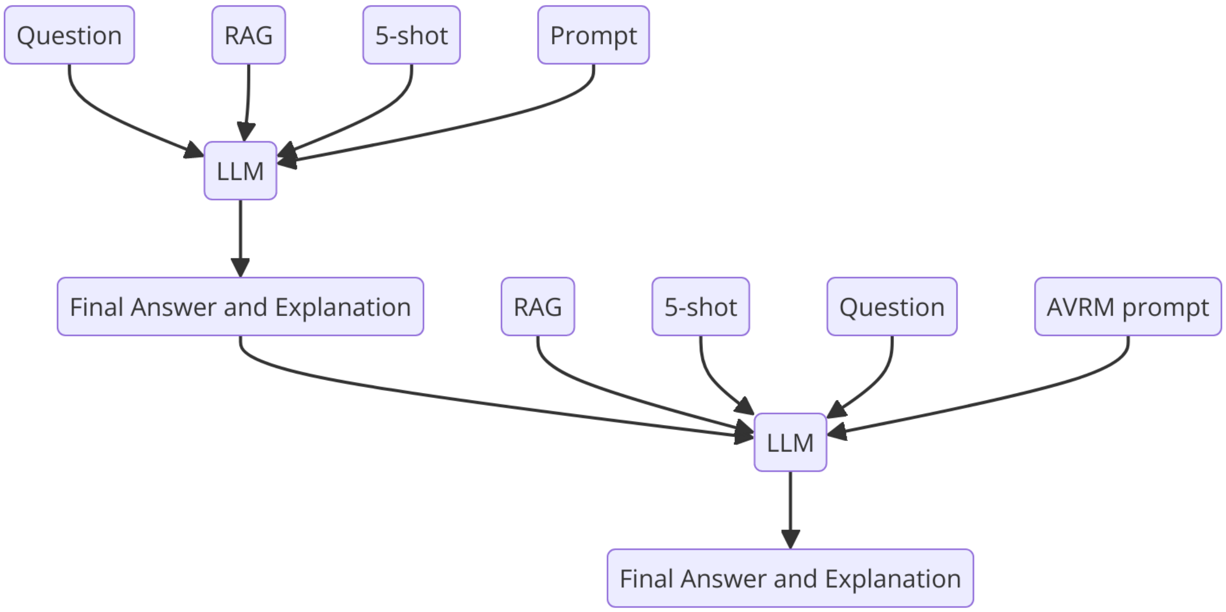


**RAG:** Retrieval Augmented Generation; **LLM:** Large Language Model.

Illustration shows the inputs and outputs of the final two steps of model prompting strategy implementation. First, we illustrate the inputs and outputs for the second to last model which utilized prompt, RAG and 5-shot to answer questions. The final model, which utilized prompt, RAG, 5-shot and AVRM received all inputs and outputs of the previous model (question, 5-shot, RAG, answer and explanation) along with an AVRM prompt to arrive at the final answer and explanation.

**Supplementary Table 1:** Example of Model Output Evaluation by the Answer Validation and Revision Model (AVRM).

| |
|---|
| **Question** |
| A 35-year-old woman presented to the emergency department with epigastric abdominal pain for 6 hours. She reveals that she is 18 weeks pregnant. The pregnancy has been uneventful thus far. She has no other past medical history. She is afebrile with normal vital signs except for a heart rate of 110 beats per minute. Laboratory test results are significant for lipase of 12,100 U/L (normal: <95 U/L), leukocyte count of 12,300/µL (normal: 4,000-10,000/µL), and bilirubin of 1.7 mg/dL (normal: 0.3-1.2 mg/dL), and otherwise normal liver chemistries. Transabdominal ultrasound confirmed the gravid uterus with a viable fetus, and cholelithiasis with a dilated common bile duct (CBD) of 8 mm. The patient was admitted to a monitored bed and intravenous fluids were administered. On the next day, her pain is significantly improved without needing analgesics. The leukocyte count and bilirubin are normal. What would be the best next step in management of this patient?<br><br>A<br>ERCP<br>B<br>EUS<br>C<br>MRCP<br>D<br>Continue observation. |
| **LLM Answer (Prompt, 5-shot, RAG)** |
| Multiple Choice Answer: D. Continue observation.<br><br>Justification: The patient presented with epigastric abdominal pain, a significantly elevated lipase level, leukocytosis, and a bilirubin level that was initially elevated, suggesting acute biliary pancreatitis likely due to gallstones. The dilation of the CBD and the presence of cholelithiasis on ultrasound supports this diagnosis. However, the patient's symptoms have improved with conservative management, her leukocyte count has normalized, and her bilirubin level has returned to normal. Given the patient's clinical improvement and the resolution of laboratory abnormalities, continued observation is appropriate at this stage.<br><br>Performing an ERCP (option A) carries a risk of post-ERCP pancreatitis, which is higher in a patient with a normal caliber bile duct and normal bilirubin. An ERCP would also expose the patient and the fetus to radiation, which should be minimized during pregnancy. EUS (option B) and MRCP (option C) are both appropriate noninvasive diagnostic alternatives to ERCP and are safe during pregnancy, as they do not involve ionizing radiation. However, both are less likely to be necessary at this point given the patient's improvement.<br><br>In this clinical scenario, as the patient's symptoms and abnormal laboratory findings have resolved with supportive care, immediate additional interventions such as ERCP, EUS, or MRCP are not indicated. It is reasonable to continue observation and reassess the need for further intervention if the patient's condition changes. Following the acute episode, management of gallstone disease in pregnancy should be supportive initially, with other interventions considered if necessary, ideally in the second trimester if the patient were to have recurrent symptoms (as explained in the additional information provided).<br><br>References:<br>- American College of Obstetricians and Gynecologists,Äô Committee on Practice Bulletins,ÄîObstetrics. |

| |
|---|
| ACOG Practice Bulletin No. 202: Gestational Hypertension and Preeclampsia. Obstet Gynecol. 2019;133(1):e1-e25.<br>- Maple JT, Ben-Menachem T, Anderson MA, et al. The role of endoscopy in the management of acute non-variceal upper GI bleeding. Gastrointest Endosc. 2012;75(6):1132-1138. |
| **LLM Answer (Prompt, 5-shot, RAG, AVRM)** |
| Multiple Choice Answer: C. MRCP.<br><br>Justification:<br>The provided answer suggests continued observation based on the improvement in clinical symptoms and normalization of laboratory values. While conservative management is often effective in cases of mild biliary pancreatitis, there is a concerning factor that should not be overlooked‚Äîthe persistent dilation of the common bile duct (CBD) as evidenced by the ultrasound findings. The diameter of the CBD at 8 mm in a pregnant woman should raise suspicion for potential biliary obstruction despite the apparent clinical improvement.<br><br>In this scenario, MRCP (Magnetic Resonance Cholangiopancreatography) would be a more prudent choice over continued observation. MRCP is a non-invasive imaging technique that provides detailed images of the bile ducts, pancreatic duct, and surrounding structures without exposing the patient or fetus to ionizing radiation. This is crucially important in the setting of pregnancy where minimizing fetal exposure to radiation is paramount. The significant dilation of the CBD, even in the absence of elevated liver chemistries at the moment, suggests an underlying obstruction possibly due to a stone or sludge which may not be adequately visualized on ultrasound alone.<br><br>Additionally, MRCP can help clarify the diagnosis without the risk of inducing pancreatitis, as might be seen with ERCP (Endoscopic Retrograde Cholangiopancreatography). Understanding the etiology of the CBD dilation is essential to guide further management, which may include endoscopic intervention if an obstructing stone is identified or continued observation if MRCP is clear.<br><br>The references provided in the answer, relevant to hypertension and non-variceal GI bleeding, do not directly support the decision-making process in this clinical scenario concerning biliary pancreatitis and pregnancy-related considerations. The management should rather align with guidelines specifically addressing hepatobiliary diseases, such as those from the American Gastroenterological Association (AGA) or the American Society for Gastrointestinal Endoscopy (ASGE). This detailed inspection and potential treatment planning via MRCP reduce the risk of a complicated course, which could prove detrimental to both the mother and the fetus if not addressed timely.<br><br>Therefore, MRCP (Option C) should be the next step in the management of this patient to accurately map the biliary anatomy and guide further therapeutic decisions. |

**RAG:** Retrieval Augmented Generation; **LLM:** Large Language Model.

**Supplementary Figure 3:** Proportion of Questions with the Most Popular Answer Chosen by GPT-4 and Gemini Pro on the 2022 ACG Self-Assessment Examination with the Implementation of Each Model Tuning Method.

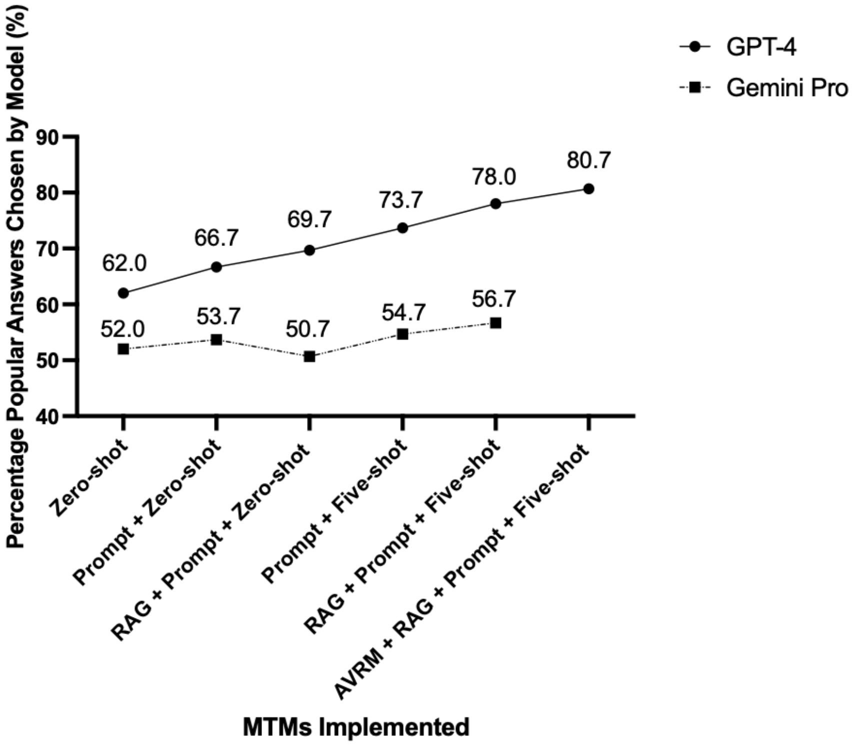


**MTM:** Model tuning methods; **GPT-4:** Generative Pre-trained Transformer 4; **ACG:** American College of Gastroenterology; **RAG:** Retrieval Augmented Generation; **AVRM:** Answer Validation Revision Model

Note: "Most Popular Answer" defined by the answer choice chosen by the largest percentage of human respondents.

We examined how frequently GPT-4 and Gemini Pro selected the most popular answer choice among human test-takers, irrespective of the correct choice (**Supplementary Figure 2**). Similar to the accuracy trends, GPT-4 outperformed Gemini Pro at all implementation steps. GPT-4's (b=4.87, p=0.001) and Gemini Pro's (b=1.51, p=0.007) performance significantly improved with the incremental addition of model tuning methods. GPT-4 showed a 3.2-fold rate of increase in improvement compared to Gemini Pro (p=0.002). Trends stratified by image and non-image-containing questions are shown in **Supplementary Figure 3**.

**Supplementary Figure 4**:  Proportion of Questions with the Most Popular Answer Chosen by GPT-4 (A) and Gemini Pro (B) on Image and Non-image containing 2022 ACG Self-Assessment Examination with the Implementation of Each Model Tuning Method.

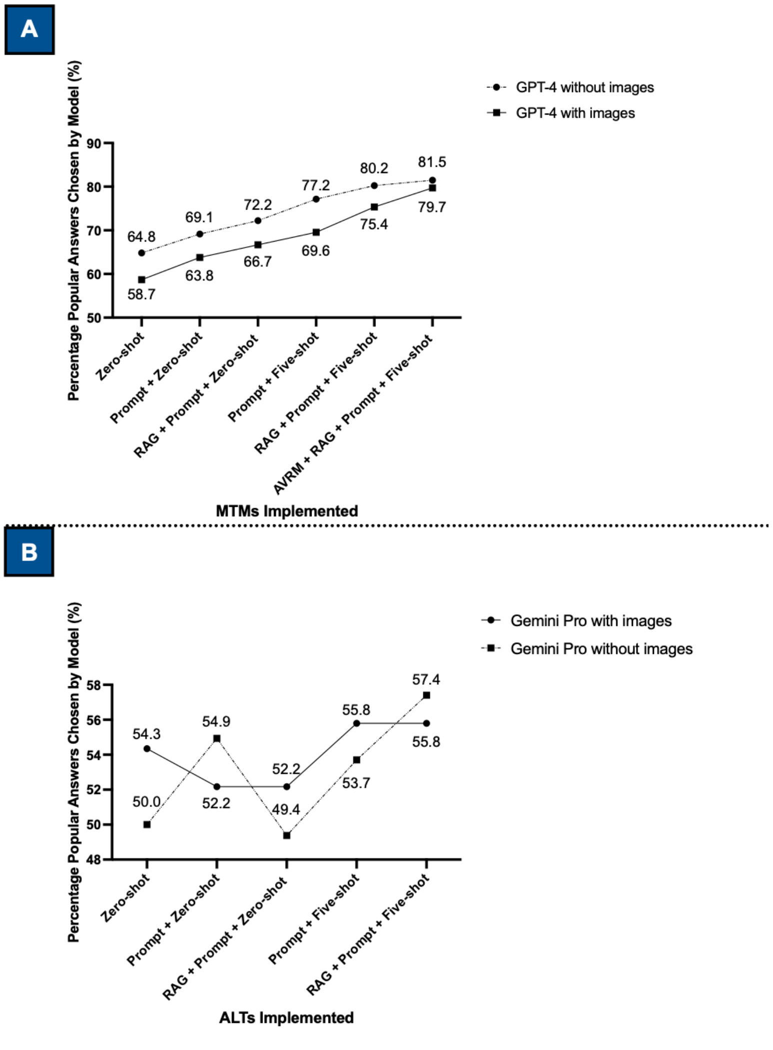


**MTM:** Model tuning methods; **GPT-4:** Generative Pre-trained Transformer 4; **ACG:** American College of Gastroenterology; **RAG:** Retrieval-Augmented Generation; **AVRM:** Answer Validation Revision Model; **ns**: no significance

Note: "Most Popular Answer" defined by the answer choice chosen by the largest percentage of human respondents.

GPT-4 demonstrated disparity in choosing the most popular answer among image and non-image-based questions when utilizing the zero-shot approach. Subsequently, GPT-4's performance significantly improved with the incremental addition of model tuning methods on both non-image (B=4.45, p-0.007) and image (B=5.36, p=0.001) based questions without a statistically significant difference among the two trends (p=0.24) (**Supplementary Figure 4A).** The most dramatic improvement in performance on image-based questions compared to non-image-based questions appears after the implementation of prompting, RAG, and 5-shot learning, followed by another relative increase in performance with the implementation of the AVRM. Gemini Pro did not demonstrate a statistically significant increase in performance with the incremental addition of model tuning methods on both non-image (B=2.10, p=0.12), and image (B=0.81, p=0.39) based questions without difference among the two trends (p=0.30) (**Figure Supplementary 4B**).

**Supplementary Figure 5**: Stepwise Performance of Gemini Pro on the 2022 ACG Self-Assessment Examination with the Implementation of Each Model Tuning Method Stratified by Question Difficulty.

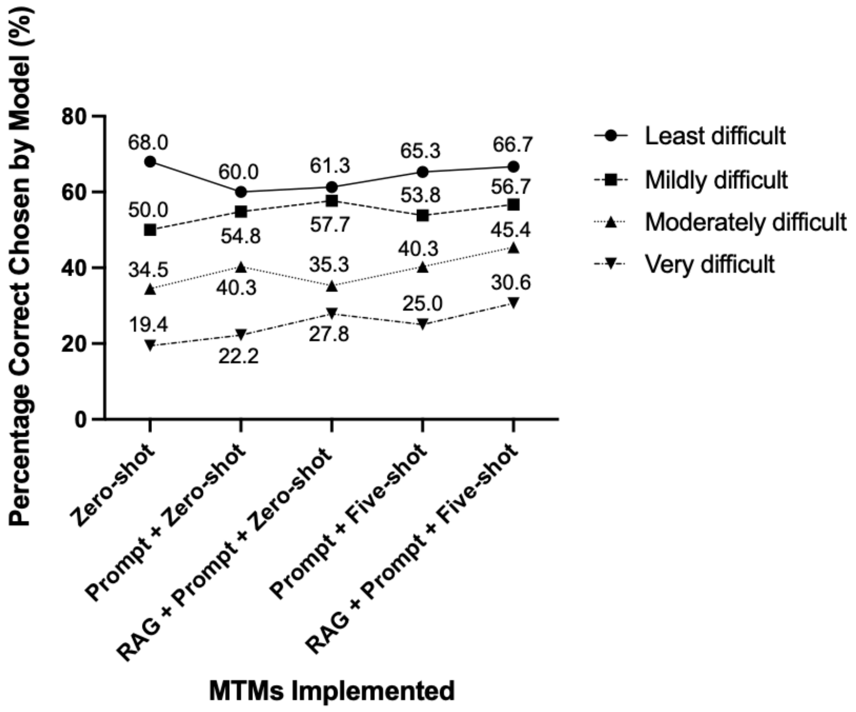


**MTM:** Model tuning methods; **ACG:** American College of Gastroenterology; **RAG:** Retrieval-Augmented Generation; **AVRM:** Answer Validation Revision Model, **ns**: no significance

Note: Question difficulty was defined by the percentage of human respondents answering a question correctly, the lower the percent accuracy, the more difficult the question.

Gemini Pro performance significantly improved with the incremental addition of model tuning methods among the very difficult (b=3.61, p=0.017) but not the least difficult (b=0.134, p=0.95), mildly difficult (b=1.92, p=0.12), or moderately difficult (b=3.28, p=0.053) questions (**Supplementary Figure 5**).

**Supplementary Figure 6**: Stepwise Performance of the Final Composite** Gemini Pro Model on the 2022 ACG Self-Assessment Examination Questions with the Implementation of Each Model Tuning Method Stratified by Topic.

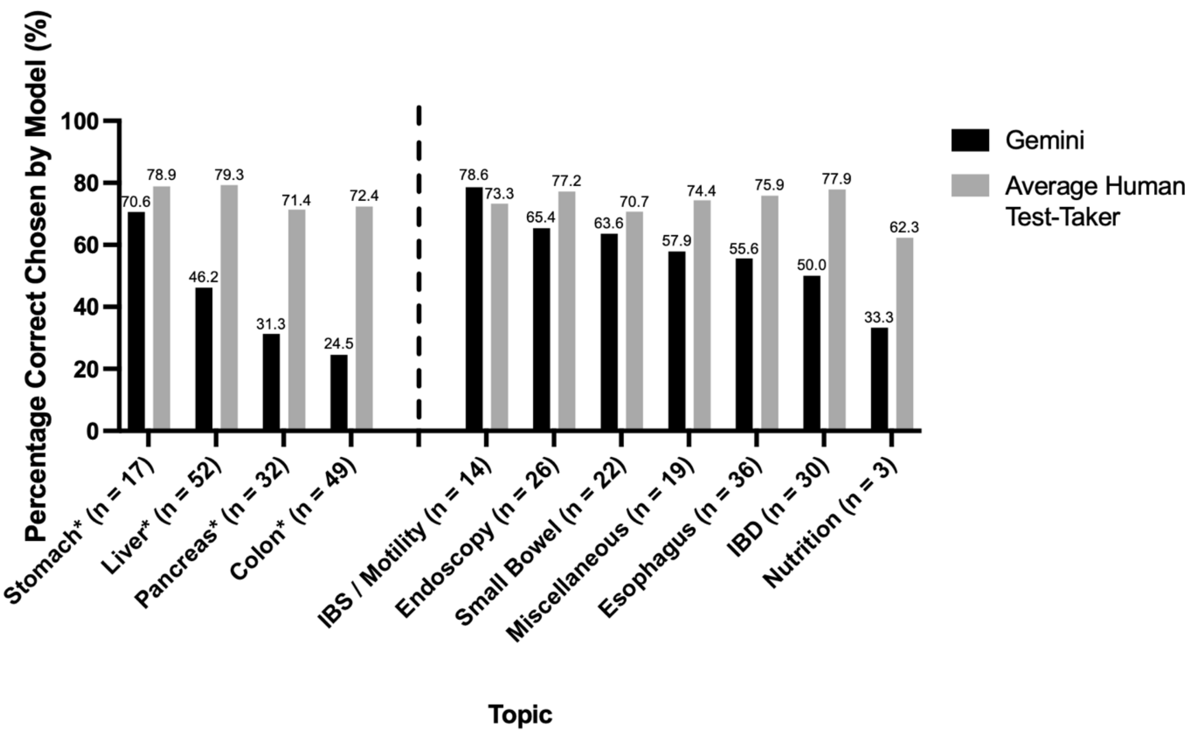


**ACG:** American College of Gastroenterology; **IBD:** inflammatory bowel disease; **IBS**: irritable bowel syndrome.

*Topics represented in 5-shot learning

**Model utilized prompt, 5-shot learning and retrieval augmented generation (RAG)

**References**

1. Recursively split by character. Accessed March 15, 2024. https://python.langchain.com/v0.1/docs/modules/data_connection/document_transformers/recursive_text_splitter/

2. Neelakantan A, Xu T, Puri R, et al. Text and Code Embeddings by Contrastive Pre-Training. Published online January 24, 2022. Accessed May 29, 2024. http://arxiv.org/abs/2201.10005
